# Timing of Regadenoson-induced Peak Hyperemia and the Effects on Coronary Flow Reserve

**DOI:** 10.1101/2024.01.15.23300449

**Authors:** Nathan Kattapuram, Shahrad Shadman, Eric E. Morgan, Charles Benton, Stacian Awojoodu, Dong-Yun Kim, Joao Ramos, Ana Barac, W. Patricia Bandettini, Peter Kellman, Gaby Weissman, Marcus Carlsson

## Abstract

**Background:** Regadenoson is used to induce hyperemia in cardiac imaging, facilitating diagnosis of ischemia and assessment of coronary flow reserve (CFR). While the regadenoson package insert recommends administration of radionuclide tracer 10-20 seconds after injection, peak hyperemia has been observed at approximately 100 seconds after injection in healthy volunteers undergoing cardiovascular magnetic resonance imaging (CMR). It is unclear when peak hyperemia occurs in a patient population.

**Objectives:** The goal of this study was to determine time to peak hyperemia after regadenoson injection in healthy volunteers and patients, and whether the recommended image timing in the package insert underestimates CFR.

**Methods:** Healthy volunteers (n=15) and patients (n=25) underwent stress CMR, including phase-contrast imaging of the coronary sinus at rest and multiple timepoints after 0.4 mg regadenoson injection. Coronary sinus flow (ml/min) was divided by resting values to yield CFR. Smoothed, time-resolved curves for CFR were generated with pointwise 95% confidence intervals.

**Results:** CFR between 60 and 120 seconds was significantly higher than CFR at 30 seconds after regadenoson injection (p < 0.05) as shown by non-overlapping 95% confidence intervals for both healthy volunteers (30 s, [2.8, 3.4]; 60 s, [3.8, 4.4]; 90 s, [4.1, 4.7]; 120 s, [3.6, 4.3]) and patients (30 s, [2.1, 2.5]; 60 s, [2.6, 3.1]; 90 s, [2.7, 3.2]; 120 s, [2.5, 3.1]).

**Conclusion:** Imaging at 90 seconds following regadenoson injection is the optimal approach to capture peak hyperemia. Imaging at 30 seconds, which is more aligned with the package insert recommendation, would yield an underestimate of CFR and confound assessment of microvascular dysfunction.

## Introduction

Stress-induced myocardial ischemia resulting from epicardial stenoses can be non-invasively diagnosed with single photon emission computed tomography (SPECT), positron emission tomography (PET), and cardiovascular magnetic resonance (CMR) imaging depicting regional perfusion defects during hyperemia that are absent at rest.^1^ The 2021 ACC/AHA guidelines for assessment of patients with stable angina and suspected ischemia with no obstructive coronary arteries (INOCA) state that quantitative stress CMR or PET with reduced coronary flow reserve (CFR), which is the ratio of stress coronary blood flow to resting flow, can be effective for assessing coronary microvascular disease.^2^ Phase-contrast magnetic resonance imaging can be used to measure flow in the coronary sinus, which drains the vast majority of left ventricular myocardium into the right atrium and is a validated technique to determine global left ventricular coronary flow and CFR.^3^ Reduced CFR has demonstrated prognostic significance for different patient cohorts.^4,5,6^

Pharmacological vasodilation is the preferred method to induce hyperemia in PET, SPECT, and CMR. Regadenoson is a selective agonist for the same A_2a_ receptor that adenosine binds to in order to mediate coronary vasodilation and has been shown to be an equivalent vasodilator to exogenous adenosine.^7^ Regadenoson is administered as a single, standard dose bolus injection compared to the continuous adenosine infusion routinely needing two venous access points. This greater ease of use has led to broader use of regadenoson instead of adenosine as the vasodilator of choice in clinical practice. Furthermore, regadenoson has shown higher tolerability and a more favorable side effect profile, particularly for patients with reactive airway disease.^8,9^

The widespread use of regadenoson and the clinical importance of CFR warrant consideration of the timing of stress imaging. Stress imaging prior to or after peak hyperemia is reached may result in an underestimation of CFR, confounding care for patients with suspected microvascular disease. A previous study demonstrated that the onset of peak regadenoson-induced hyperemia in healthy subjects occurs later than the time suggested in the regadenoson package insert,^10^ which recommends administration of radionuclide tracer 10-20 seconds after regadenoson injection for PET and SPECT imaging.^11^While regadenoson has received FDA approval for nuclear imaging, the use of regadenoson in CMR stress perfusion imaging in the USA is technically off-label. The time course of the regadenoson-induced hyperemic response has not previously been investigated in patients using CMR. Therefore, the aim of this study was to determine the time to peak regadenoson-induced hyperemia in patients and healthy volunteers (HV) and whether that time corresponds with the regadenoson package insert recommendation for image timing during hyperemia.

## Methods

### Study Population

After signing the informed consent, seventeen adult HV and 29 patients, with either clinical or research indications, underwent CMR with regadenoson injection. Exclusion criteria for both cohorts included contraindications to CMR or regadenoson and hypersensitivity to gadolinium-based contrast. Exclusion criteria for HV included history of any known major cardiovascular risk factors including hypertension, diabetes, familial hyperlipidemia, angina, dyspnea, myocardial infarction, cerebrovascular or peripheral vascular disease, structural or functional cardiac impairment, or current smoking. Subjects were instructed to abstain from caffeine for 24 hours prior to the time of their scan as caffeine is known to blunt adenosine receptor-mediated hyperemia.^12^ Serum caffeine levels were checked to confirm sufficient caffeine abstinence, and subjects with serum caffeine levels exceeding 5 µg/ml, the threshold at which caffeine has been shown to significantly blunt CFR, were excluded from analysis.^13^ Patients were not asked to refrain from taking prescribed medications prior to scanning. The study was approved by the National Institutes of Health Institutional Review Board (IRB # 000299).

### CMR Imaging

CMR was conducted using a 1.5T scanner (Magnetom Sola, Siemens AG, Erlangen, Germany) with an 18-channel receive coil and 32-channel spinal coil. Short-axis and long-axis cine and late gadolinium enhancement (LGE) imaging were acquired. The coronary sinus was identified on axial and short-axis localizers. The plane for phase-contrast imaging was oriented perpendicular to the direction of flow in the coronary sinus, approximately 5-10 mm from the ostia. Coronary sinus flow (ml/min) was measured with 2D phase-contrast cine imaging of the coronary sinus at rest and at multiple timepoints after regadenoson injection. First-pass perfusion imaging was only conducted for patients. A summary of the imaging protocol for patients and HV, including the timepoints at which the coronary sinus was imaged, is shown in Figure 1.

**Figure 1:**
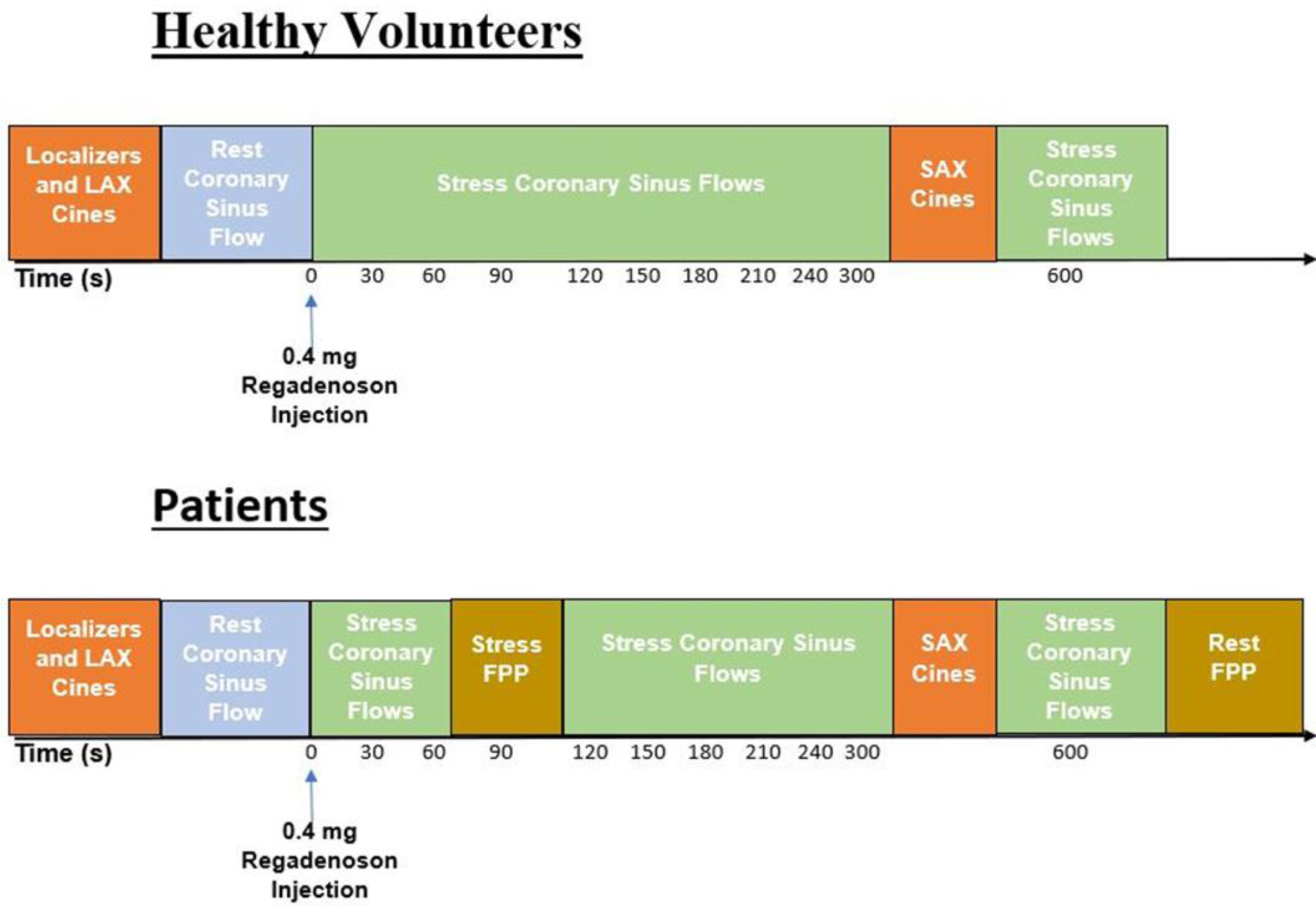
Imaging Protocol. Following initiation of scan, localizers and long-axis (LAX) cines were collected. Phase-contrast imaging of the coronary sinus was acquired before and at repeated timepoints after regadenoson injection. Only patients had first-pass perfusion (FPP) imaging at stress, with acquisition 90 seconds after regadenoson injection, and rest. Therefore, patients did not have coronary sinus flow imaging at 90 seconds after regadenoson injection. Short-axis (SAX) cine imaging was acquired for determination of left ventricular volumes, function and mass. FPP = First-pass perfusion; LAX = Long-axis; SAX = Short-axis.

Sequence parameters were as follows: Steady-state free precession (SSFP) cine images (Echo time (TE): 1.1 ms; flip angle: 54 degrees; acquired temporal resolution: 28 ms; in-plane spatial resolution: 2.0 mm; slice thickness: 8 mm; slice gap: 0 mm), first-pass SSFP perfusion (TE: 1.2 ms; flip angle: 12 degrees; acquired temporal resolution: 188.2 ms; in-plane spatial resolution: 2.25 mm; slice thickness: 8 mm; inversion time: 105 ms), motion-corrected phase-sensitive inversion recovery SSFP LGE (TE: 1.1 ms; flip angle: 55 degrees; in-plane spatial resolution: 2 mm; slice thickness: 8 mm), and 2D phase-contrast cine imaging of the coronary sinus (TE: 2.3 ms; flip angle: 15 degrees; acquired temporal resolution: 50 ms; in-plane spatial resolution: 2.0 mm; slice thickness: 8 mm). Velocity encoding of the 2D phase-contrast cine imaging of the coronary sinus was 80 and 120 cm/s for acquisitions before regadenoson injection and exclusively 120 cm/s for acquisitions after injection.

Regadenoson was administered as a 0.4 mg bolus injection over 10 seconds, followed by 5 mL saline flush under continuous monitoring of MRI compatible electrocardiography. Heart rate and systolic blood pressure (SBP) were recorded in subjects before regadenoson injection. SBP was also recorded approximately two minutes after regadenoson injection and during post-stress recovery. Mean heart rate during phase-contrast imaging of the coronary sinus at rest and at multiple timepoints after regadenoson was recorded to evaluate regadenoson-induced changes in heart rate.

### Image Analysis

Left ventricular planimetry was performed with machine learning algorithms available in Segment CMR (Segment CMR version 4.0 R10221, Medviso, Lund, Sweden) to yield ventricular volumes and left ventricular mass.^14^ Manual adjustments were made to epicardial and endocardial contours as needed. The coronary sinus in the 2D phase-contrast cine images was also delineated automatically and manually corrected as needed, with guidance from both the phase and magnitude images. Coronary sinus delineations were propagated across the cardiac cycle using a validated automatic algorithm in Segment CMR to yield flow vs time curves and determine the rate of coronary sinus flow (ml/minute).^15^ Background correction was not applied. Coronary sinus flow was divided by left ventricular mass (g) to yield perfusion (ml/min/g). CFR was calculated at multiple timepoints for each subject by dividing each coronary sinus flow measurement after regadenoson injection by the subject’s resting coronary sinus flow. If a subject had multiple resting coronary sinus flow acquisitions with adequate image quality with velocity encoding at 80 cm/s and 120 cm/s, the average of the two measurements was used to determine resting coronary sinus flow.

An experienced reader visually assessed first-pass perfusion and LGE images. Regional inducible ischemia was assessed if there was a perfusion defect present during hyperemia that was not present at rest and that was more extensive than potential LGE findings in that segment. LGE findings were classified as ischemic or non-ischemic pattern.

### Statistical Analysis

For continuous variables, mean differences between HV and patients were analyzed using two-sample t tests. For categorical variables, the homogeneity of distribution was tested using Fisher’s exact test. All statistical analysis was performed using GraphPad Prism (version 9.0.2 for Windows, GraphPad Software, Boston, Massachusetts USA) and R (version 4.0 or later, The R Foundation). Due to the exploratory nature of this study, no adjustment for multiple tests was made. As there was not a uniform set of timepoints for coronary sinus flow measurements between subjects, Lowess smoothing was used to generate interpolated time series for both heart rate and CFR and estimate values between timepoints used for measurement. Smoothed time-resolved curves for stress to rest heart rate ratio and CFR were generated with pointwise 95% confidence intervals at 30 second intervals between 30 seconds and 240 seconds after regadenoson injection and the 300 second timepoint (Figs. 2,3). Data for the time interval between 600 and 720 seconds are shown as a single time point. Unless otherwise stated, continuous variables are reported as mean ± standard deviation.

**Figure 2:**
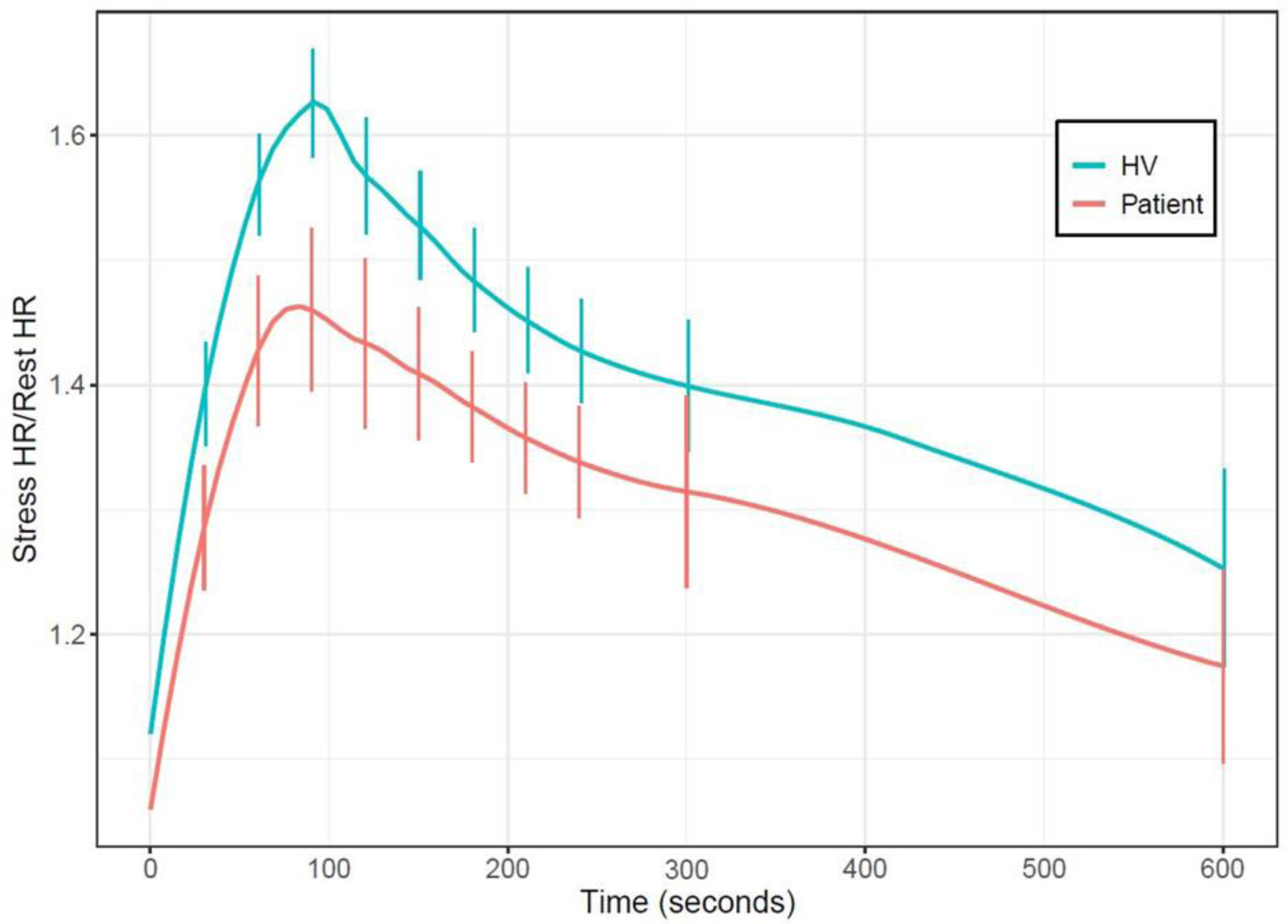
Time Course of Relative Heart Rate Response to Regadenoson. Smoothed time-resolved curves for the ratio between stress heart rate and rest heart rate following regadenoson injection for patient and HV cohorts, along with 95% confidence intervals at various points. Stress to rest heart rate ratio measured between 10 and 12 minutes after injection is collapsed and shown at the 600 second timepoint. HV = Healthy volunteers.

## Results

### Cohort Characteristics

Among the HV, 6% experienced chest discomfort and flushing respectively with regadenoson stress. Among the patients, 7% experienced dyspnea, 3% had chest discomfort, and 7% reported flushing with regadenoson stress. Two HV and 2 patients were retrospectively excluded due to inadequate coronary sinus image quality preventing CFR calculation, and one patient was excluded after having incomplete stress imaging due to dyspnea. One additional patient had serum caffeine levels exceeding 5 µg/ml and was excluded from analysis. Characteristics of the 15 HV and 25 patients included in the analysis are shown in Table 1. One analyzed patient underwent coronary sinus imaging at rest and stress before requesting premature termination of the scan before acquisition of short-axis cines, thus preventing quantitative assessment of the subject’s ventricular function and mass. Patients were older (64 ± 17 years) than HV (43 ± 13 years, *p*=0.001), and the majority of subjects were women (HV 73% and patients 56%, *p*=0.329).

**Table 1.** Cohort Characteristics.

|  | **HV** | **Patients** | ***p*** |
| --- | --- | --- | --- |
| **Demographics** |  |  |  |
| n | 15 | 25 |  |
| Age | 43 ± 13 | 64 ± 17 | <0.001 |
| Sex-female (%) | 11 (73) | 14 (56) | 0.329 |
| **Medical History** |  |  |  |
| Chest pain (%) |  | 5 (20) |  |
| Hypertension (%) |  | 12 (48) |  |
| Hyperlipidemia (%) |  | 19 (76) |  |
| T2DM (%) |  | 4 (16) |  |
| Heart Failure (%) |  | 7 (28) |  |
| Myocardial Infarction (%) |  | 5 (20) |  |
| Prior PCI (%) |  | 5 (20) |  |
| **Medications of Interest** |  |  |  |
| β-blockers |  | 8 (32) |  |
| Ca-channel antagonists |  | 6 (24) |  |
| Oral Nitrates |  | 5 (20) |  |
| **MRI Findings** |  |  |  |
| LVEF (%) | 70 ± 5 | 60 ± 12 | 0.004 |
| LVEDVI (ml/m<sup>2</sup>) | 70 ± 12 | 68 ± 16 | 0.713 |
| LVESVI (ml/m<sup>2</sup>) | 21 ± 6 | 27 ± 11 | 0.049 |
| SVI (ml/m<sup>2</sup>) | 49 ± 8 | 41 ± 11 | 0.023 |
| LVMI (g/m<sup>2</sup>) | 51 ± 9 | 58 ± 14 | 0.067 |
| First-Pass Perfusion Imaging showing Myocardial Ischemia (%) |  | 8 (32) |  |

Values are presented as mean ± standard deviation. HV= Healthy volunteers; LVEDVI=Left
ventricular end-diastolic volume index; LVEF= Left ventricular ejection fraction; LVESVI=Left
ventricular end-systolic volume index; LVMI=Left ventricular mass index; PCI = Percutaneous
coronary intervention; SVI=Stroke volume index; T2DM = Type 2 Diabetes mellitus.

Hypertension, hyperlipidemia, and type 2 diabetes mellitus were present in 48%, 76%, and 16% of patients respectively. Five patients had history of myocardial infarction and prior percutaneous coronary intervention. Seven patients had clinical diagnosis of heart failure. With regard to medications at the time of study, 20%, 24%, and 32% of patients had prescriptions for oral nitrates, calcium channel antagonists, and beta blockers respectively. Left ventricular ejection fraction (LVEF) was lower in patients compared to HV (60 ± 12% vs 70 ± 5%, *p*=0.004). Twelve patients exhibited LGE (3 with ischemic and 9 with non-ischemic pattern), and eight were considered to have myocardial ischemia on first-pass perfusion.

### Hemodynamic Response to Regadenoson

The resting heart rates of HV and patients were not significantly different (HV 63 ± 7, Patients 68 ± 11, *p=*0.134). However, after regadenoson injection, HV had higher increases in heart rate, and in turn, a higher mean peak stress to rest heart rate ratio compared to patients (1.7 ± 0.2 vs 1.5 ± 0.2, *p*=0.026). Resting and stress SBP of patients were not significantly different from the corresponding values in HV (Resting SBP: HV 118 ± 10 mmHg, Patients 125 ± 18 mmHg, *p*= 0.175; Stress SBP: HV 125 ± 12 mmHg, Patients 122 ± 21 mmHg, *p*=0.672) (Table 2). Patients and HV had similar times to peak heart rate following regadenoson injection (Figure 2).

**Table 2.**
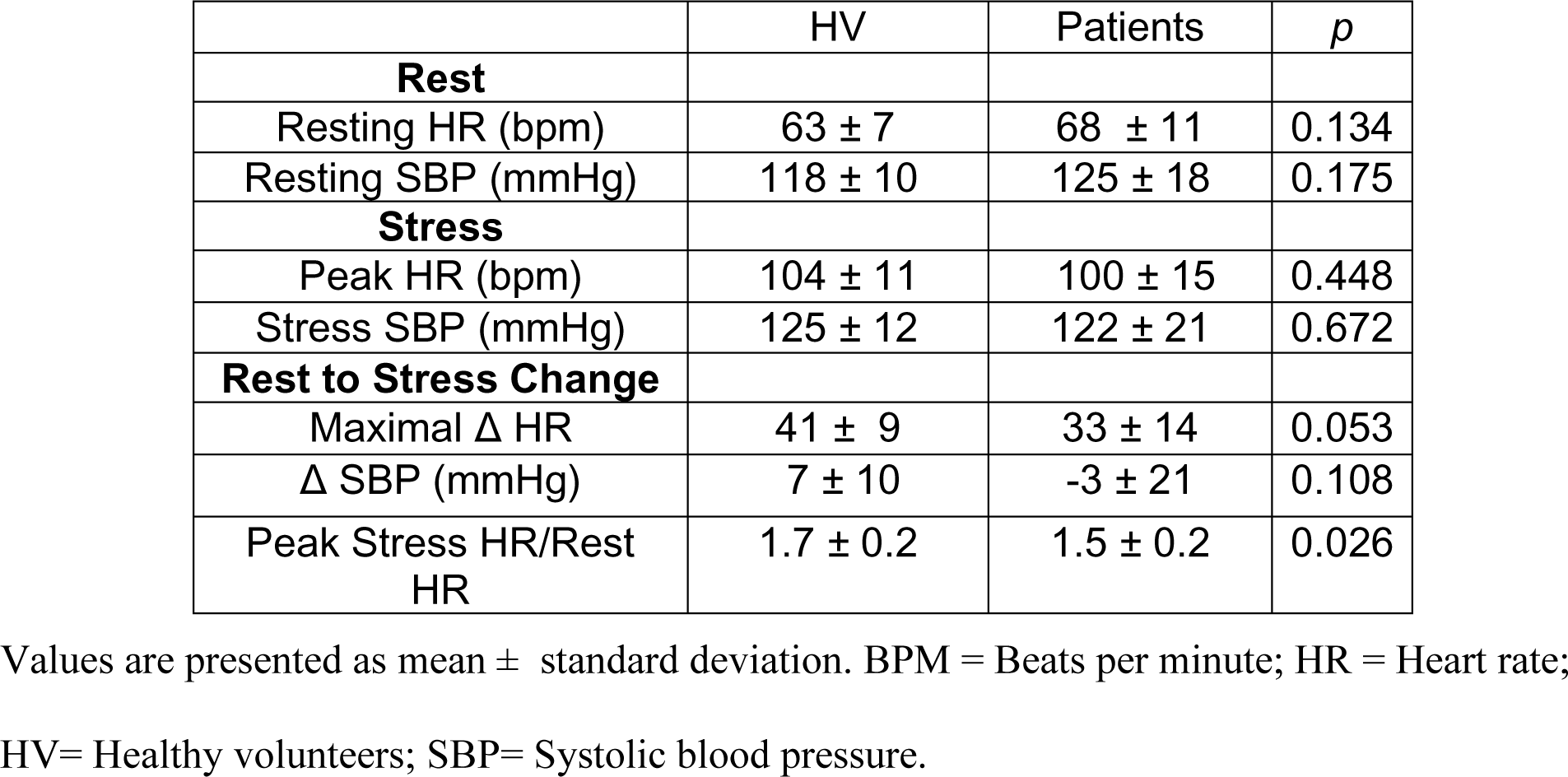
Hemodynamic Characteristics.

### Hyperemic Response to Regadenoson

Resting coronary sinus flow and perfusion were not significantly different between patients and HV (Table 3). However, following regadenoson injection, patients had lower peak coronary sinus flow (411 ± 100 ml/min vs 519 ± 132 ml/min, p=0.006) and perfusion (4.0 ± 1.1 ml/min/g vs 5.4 ± 1.4 ml/min/g, p=0.001) than HV.

**Table 3.** Perfusion Metrics.

|  | HV | Patients | *p* |
| --- | --- | --- | --- |
| **Rest** |  |  |  |
| Resting HR (bpm) | 63 ± 7 | 68 ± 11 | 0.134 |
| Resting Coronary Sinus Flow (ml/min) | 109 ± 25 | 137 ± 50 | 0.051 |
| Resting Perfusion (ml/min/g) | 1.1 ± 0.2 | 1.3 ± 0.5 | 0.234 |
| **Stress** |  |  |  |
| Peak HR (bpm) | 104 ± 11 | 100 ± 15 | 0.448 |
| Peak Coronary Sinus Flow (ml/min) | 519 ± 132 | 411 ± 100 | 0.006 |
| Peak Perfusion (ml/min/g) | 5.4 ± 1.4 | 4.0 ± 1.1 | 0.001 |
| **Rest to Stress Change** |  |  |  |
| Peak CFR | 4.9 ± 1.5 | 3.3 ± 1.1 | <0.001 |

Perfusion metrics derived from coronary sinus imaging. Coronary flow reserve (CFR) was calculated as the ratio between stress coronary sinus flow and rest flow. Values presented as mean ± standard deviation. HR= Heart rate; HV= Healthy volunteers.

The interpolated time course for CFR for both cohorts shows the onset of peak CFR at 90 seconds after regadenoson injection (Central Illustration). As indicated by the non-overlapping 95% confidence intervals, CFR values at 60, 90, and 120 seconds after regadenoson injection were each significantly higher than CFR values at 30 seconds (p<0.05) in both patients (30 s, [2.1, 2.5]; 60 s, [2.6, 3.1]; 90 s, [2.7, 3.2]; 120 s, [2.5, 3.1]) and HV (30 s, [2.8, 3.4]; 60 s, [3.8, 4.4]; 90 s, [4.1, 4.7]; 120 s, [3.6, 4.3]).

## Discussion

This study demonstrates that peak hyperemia occurs at approximately 90 seconds after regadenoson injection, and that CFR measured between 60 and 120 seconds is elevated compared to CFR measured at 30 seconds. Our results have implications for myocardial perfusion imaging regardless of modality when using regadenoson for both detection of regional ischemia and microvascular dysfunction. The regadenoson package insert recommendation would result in underestimation of CFR and overestimation of microvascular disease.

### Considerations of Image Timing in Calculation of CFR

The recommendation in the package insert of regadenoson is derived from previous study results which found the onset of mean near peak regadenoson-induced hyperemia to be 33 seconds after injection, independent of dose in patients undergoing clinically indicated cardiac catheterization.^16^ The explanation for different findings of the time for peak hyperemia between the study by Lieu et al. and ours is not evident. There are methodological differences between the study that informed the package insert recommendation, which used catheter-based peak velocity measurements (cm/s) in either the left anterior descending or the left circumflex artery to assess hyperemia,^16^ and this study, which measured peak cardiac venous flow (ml/min). The catheter-based velocity measurements do not account for changes in vessel diameter, while the 2D phase-contrast cine CMR coronary sinus flow measurements account for all flow velocities within the vessel cross-sectional area. Increases in vessel diameter after the time at which peak velocity is reached may explain why this study finds the onset of peak hyperemia later than the study informing the package insert recommendation. The other difference is measuring proximal (catheter-based in coronary artery) or distal (MRI in coronary sinus) to the microvascular bed. The hyperemia is caused by the decreased resistance in the microvascular bed, and the effect will be an almost simultaneous increase of flow proximal and distal to the microvasculature. Otherwise, there would be a stasis of blood in the myocardium. Therefore, it is unlikely that the different anatomical places of measurement explain the difference of roughly 60 seconds between the study referenced in the package insert and this study.

Interpolated representations show that CFR at 30 seconds after regadenoson injection, which aligns with the package insert recommendation, is significantly lower than CFR between 60 and 120 seconds after regadenoson injection respectively for both cohorts. Thus, the regadenoson package insert recommendation would result in the underestimation of CFR and incorrect overdiagnosis of microvascular disease. Stress imaging between 60 and 120 seconds and ideally at 90 seconds after regadenoson injection would be a superior approach to capture peak hyperemia for the accurate calculation of CFR.

Of note, the patients and HV both exhibited residual hyperemia up to 600 seconds after regadenoson injection, a phenomenon that has been observed at even 15 minutes after regadenoson injection and is likely attributed to the long terminal half-life of the compound.^17,18^ These findings point to the risk of conducting rest perfusion imaging within the 10 minutes following regadenoson injection, which may result in an underestimate of CFR. Residual hyperemia can be resolved with administration of aminophylline; however aminophylline administration has been shown to be unsuccessful in reversing the heart rate increase associated with regadenoson administration.^16^ This study acquired resting coronary sinus flow images prior to stress flow images, which is a clinically useful way to determine CFR.

### Considerations of Image Timing in Calculation of Stress First-Pass Perfusion Imaging

In first-pass perfusion imaging during regadenoson stress, the gadolinium-based contrast agent should ideally be administered when the patient reaches peak coronary hyperemia to accentuate differences in flow between coronary territories perfused by diseased and non-pathological vessels. When considering the potential clinical impact of conducting stress first-pass perfusion imaging before peak hyperemia is reached, it is useful to draw an analogy between premature first-pass perfusion imaging after regadenoson injection and first-pass perfusion imaging in a patient with elevated caffeine. Previous research shows that a caffeine abstinence period of 12 hours was associated with lower CFR measurements in adenosine stress CMR compared to those taken in the same healthy subjects after a caffeine abstinence period of 24 hours.^12^ The administration of caffeine, which blunts hyperemia mediated by adenosine and regadenoson, has also been shown to reduce perfusion defect size in patients with known or suspected coronary artery disease who underwent SPECT myocardial perfusion imaging during regadenoson stress.^19^ If imaging in a state of caffeine-induced submaximal hyperemia yields underestimates of perfusion defect size, we would infer that stress first-pass perfusion imaging in a state of submaximal hyperemia due to premature image timing would also have reduced sensitivity for the full extent of perfusion defects.

### Limitations

Our study was conducted with a relatively small sample size of both patients and HV, which may have obscured cohort-level differences in kinetics of peak hyperemia. We included a mixed patient population where some had previous diagnoses of heart failure and myocardial infarction. Prior studies have shown that patients with severe left ventricular systolic dysfunction required a higher dose of intravenous adenosine infusion (210 ug/kg/min) to yield maximal hyperemia while the standard adenosine dose (140 ug/kg/min) was sufficient for healthy subjects and patients with LVEF above 40%.^20^ If severity of systolic dysfunction correlates with altered pharmacodynamics of A_2a_ receptor agonists, it is possible that studying a patient cohort with a greater heart failure burden than the one featured in this study may highlight differences in the time series of the perfusion response to regadenoson compared to HV. Patient and HV cohorts were not age-matched in this study. However, the aim of this study was to determine time to peak hyperemia rather than the magnitude of hyperemia. As HV and patients had similar time courses after regadenoson injection, the age disparity between the cohorts does not detract from this study’s conclusions.

As noted previously, there was not a fully uniform set of timepoints for flow measurements. Uneven sampling of the time series may have resulted in missing peak or near peak CFR measurements and reduced the accuracy of the results. However, the use of interpolated graphs offsets the uneven sampling of the time series between patients and HV.

## Conclusion

Imaging at approximately 90 seconds after regadenoson injection is an optimal approach for capturing peak hyperemia, despite being later compared to what is suggested in the regadenoson package insert. Imaging at 30 seconds after regadenoson injection, which is in line with the package insert recommendation would result in underestimation of CFR, overestimation of vasodilatory impairment, and have reduced specificity for microvascular disease.

## Data Availability

All data produced in the present study are available upon reasonable request to the authors.

## Abbreviations

CFR: Coronary flow reserve
CMR: Cardiovascular magnetic resonance
HV: Healthy volunteers
LGE: Late gadolinium enhancement
LVEF: Left ventricular ejection fraction
PET: Positron emission tomography
SBP: Systolic blood pressure
SPECT: Single photon emission computed tomography
SSFP: Steady-state free precession
TE: Echo time

## Acknowledgements

Central illustration created with BioRender.com.

## Author’s Contributions

M.C., P.K. and W.P.B. conceived the study. M.C., A.B. and G.W. designed the study. C.B performed image acquisition. N.K, E.E.M, S.S, C.B, S.A, and J.R collected patient data and imaging data. N.K interpreted data and drafted manuscript and all authors thereafter provided critical content revisions. N.K and D.K performed statistical analysis. E.E.M, G.W., A.B and M.C provided experienced imaging guidance and assessment of patient data and images. All authors read and approved the manuscript.

## Availability of Data and Materials

The datasets used and/or analyzed during the current study are available from the corresponding author on reasonable request.

## Central Illustration

**Central Illustration:**
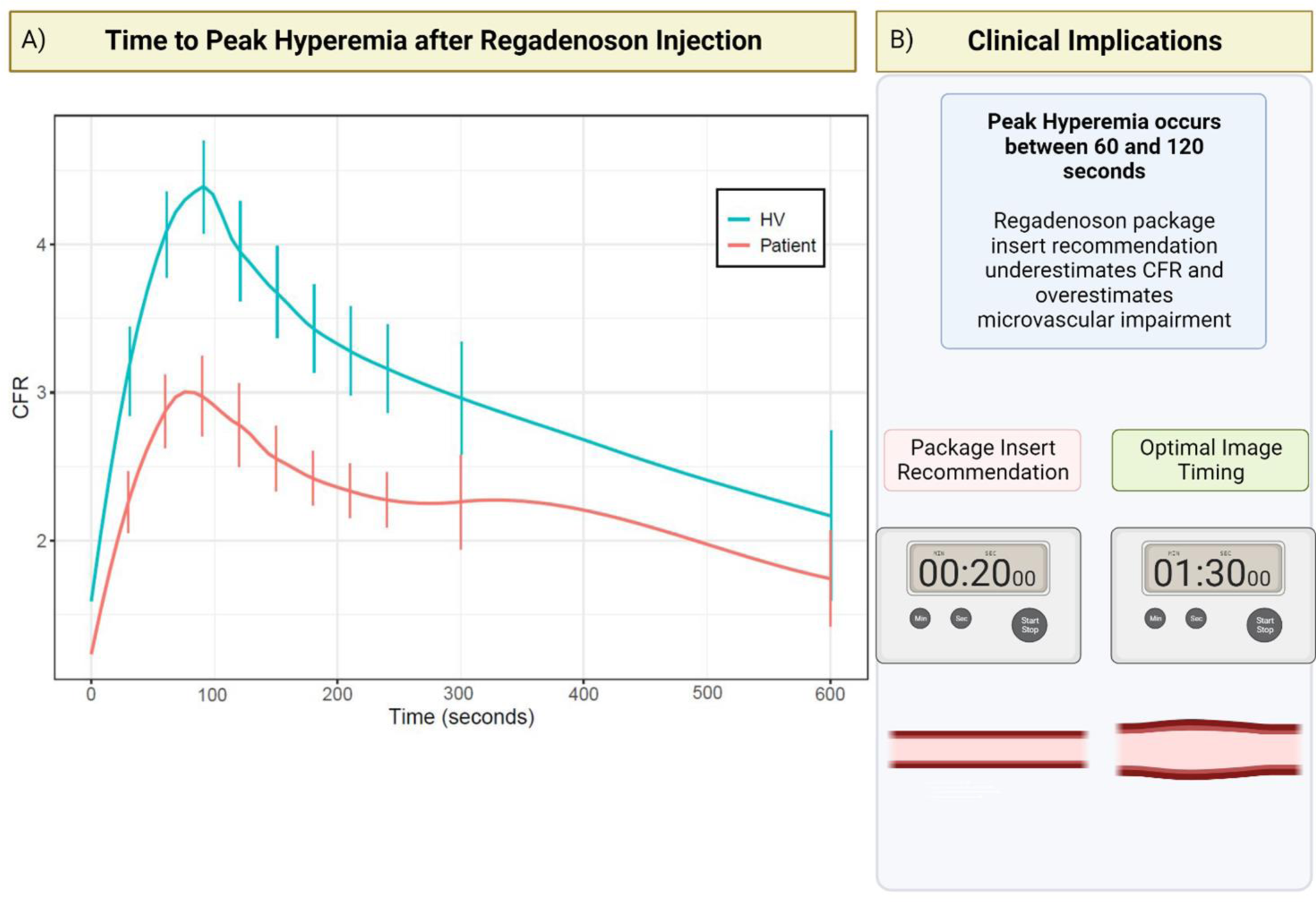
Time Course of Regadenoson-induced Hyperemia and Implications for CFR. A) Smoothed time-resolved curves for CFR following regadenoson injection for patient and HV groups, along with 95% confidence intervals at various timepoints. CFR measured between 10 and 12 minutes after injection is collapsed and shown at the 600 second timepoint. B) CFR between 60 and 120 seconds is higher CFR at 30 seconds at regadenoson injection. Imaging at 90 seconds is optimal as opposed to imaging at the earlier timepoint corresponding with the regadenoson package insert recommendation, which calls for administration of radionuclide tracer 10-20 seconds after regadenoson injection. HV = Healthy volunteers.

